# Age profile of susceptibility, mixing, and social distancing shape the dynamics of the novel coronavirus disease 2019 outbreak in China

**DOI:** 10.1101/2020.03.19.20039107

**Authors:** Juanjuan Zhang, Maria Litvinova, Yuxia Liang, Yan Wang, Wei Wang, Shanlu Zhao, Qianhui Wu, Stefano Merler, Cecile Viboud, Alessandro Vespignani, Marco Ajelli, Hongjie Yu

**Affiliations:** School of Public Health, Fudan University, Key Laboratory of Public Health Safety, Ministry of Education, Shanghai, China; ISI Foundation, Turin, Italy; Hunan Provincial Center for Disease Control and Prevention, Changsha, China; Bruno Kessler Foundation, Trento, Italy; Division of International Epidemiology and Population Studies, Fogarty International Center, National Institutes of Health, Bethesda, MD, USA; Laboratory for the Modeling of Biological and Socio-technical Systems, Northeastern University, Boston, MA USA

## Abstract

Strict interventions were successful to control the novel coronavirus (COVID-19) outbreak in China. As transmission intensifies in other countries, the interplay between age, contact patterns, social distancing, susceptibility to infection and disease, and COVID-19 dynamics remains unclear. To answer these questions, we analyze contact surveys data for Wuhan and Shanghai before and during the outbreak and contact tracing information from Hunan Province. Daily contacts were reduced 7-9 fold during the COVID-19 social distancing period, with most interactions restricted to the household. Children 0-14 years were 59% (95% CI 7-82%) less susceptible than individuals 65 years and over. A transmission model calibrated against these data indicates that social distancing alone, as implemented in China during the outbreak, is sufficient to control COVID-19. While proactive school closures cannot interrupt transmission on their own, they reduce peak incidence by half and delay the epidemic. These findings can help guide global intervention policies.

---

The novel coronavirus disease 2019 (COVID-19) epidemic caused by SARS-CoV-2 originated in Wuhan City, China in December 2019 and quickly spread nationally and globally, with 179,111 cases reported in 159 countries/territories/areas as of March 17, 2020 (*1*). A total of 80,894 cases of COVID-19, including 3,237 deaths, have been reported in mainland China, including 50,005 cases in Wuhan City and 361 cases in Shanghai City. The epidemic in Wuhan and in the rest of China slowed down after implementation of strict containment measures and movement restrictions (*2*). However key questions remain about the age profile of susceptibility, symptoms, and infectivity with this disease, how social distancing alters age-specific contact patterns, and how these factors interact to affect transmission. These questions will have a profound effect on the choice of control policies in locations where COVID-19 transmission is now intensifying. In this study, we evaluate changes in human mixing patterns brought about by social distancing by conducting contact surveys in the midst of the epidemic in two Wuhan and Shanghai. We also estimate age differences in susceptibility to infection and clinical disease based on contact tracing information gathered by the Hunan Provincial Center for Disease Control and Prevention (CDC), China. Based on those findings, we develop a mathematical transmission model to disentangle how transmission is affected by age differences in the biology of COVID-19 infection and disease, and altered mixing patterns due to social distancing. In turn, we project the impact of social distancing and school closure on COVID-19 transmission.

To estimate changes in age-mixing patterns associated with COVID-19 interventions, we performed contact surveys in two cities, Wuhan, the epicenter of the outbreak, and Shanghai, one of the largest and most densely populated cities in southeast China. Shanghai experienced extensive importation of COVID-19 cases from Wuhan as well as local transmission(*3*). The surveys were conducted from February 1, 2020 to February 10, 2020, as transmission of COVID-19 peaked across China and stringent interventions were in place. Participants in Wuhan were asked to complete a questionnaire describing their contact behavior (*4, 5*) on two different days: i) a regular weekday between December 24, 2020 and December 30, 2020, before the COVID-19 outbreak was officially recognized by the Wuhan Municipal Health Commission (used as baseline); and ii) the day before the interview (outbreak period). A similar survey was conducted in Shanghai to obtain information on contacts during the COVID-19 outbreak period; contacts for the baseline period were based on a survey using the same design conducted in the same city in 2017-2018 (*6*). Details are given in the Supplementary Material.

We analyzed a total of 1,245 contacts reported by 636 study participants in Wuhan, and 1,296 contacts reported by 557 participants in Shanghai. In Wuhan, the average daily number of contacts per participant was significantly reduced from 14.6 for a regular weekday (weighted mean contacts by age structure: 14.0) to 2.0 for the outbreak period (weighted mean contacts by age structure: 1.9) (p<0.001). The reduction in contacts was significant for all stratifications by sex, age group, type of profession, and household size, except for pre-school children aged 0-6 years old (Tab. 1). A larger reduction was observed in Shanghai, where the average daily number of contacts declined from 20.6 (weighted mean contacts by age structure: 21.7) to 2.3 (weighted mean contacts by age structure: 2.1). Although an average individual in Shanghai reported more contacts than one in Wuhan on a regular weekday, this difference disappeared during the COVID-19 outbreak period.

The typical features of age-mixing patterns(*5, 6*) emerge in Wuhan and Shanghai when we consider the regular baseline weekday period (Fig. 1A and 1D). These features can be illustrated in the form of age-stratified contact matrices (provided as ready-to-use tables in the Supplementary Materials), where each cell represents the average number of contacts that an individual has with other individuals, stratified by age groups. The bottom left corner of the matrix, corresponding to contacts between school age children, is where the largest number of contacts is recorded. The contribution of contacts in the workplace is visible in the central part of the matrix, while the three diagonals (from bottom left to top right) represent contacts between household members. In contrast, for the outbreak period where strict social distancing was in place, much of the above-mentioned features disappears, essentially leaving the sole contribution of household mixing (Fig. 1B and 1E). In particular, contacts between school-age individuals are fully removed, as highlighted by differencing baseline and outbreak matrices (Fig. 1C and 1F). Overall, contacts during the outbreak mostly occurred at home with household members (94.1% in Wuhan and 78.5% in Shanghai), thus the outbreak contact matrix nearly coincides with the within-household contact matrix in both study sites and the assortativity by age feature observed for regular days almost entirely disappear (see Supplementary Materials).

**Figure 1.**
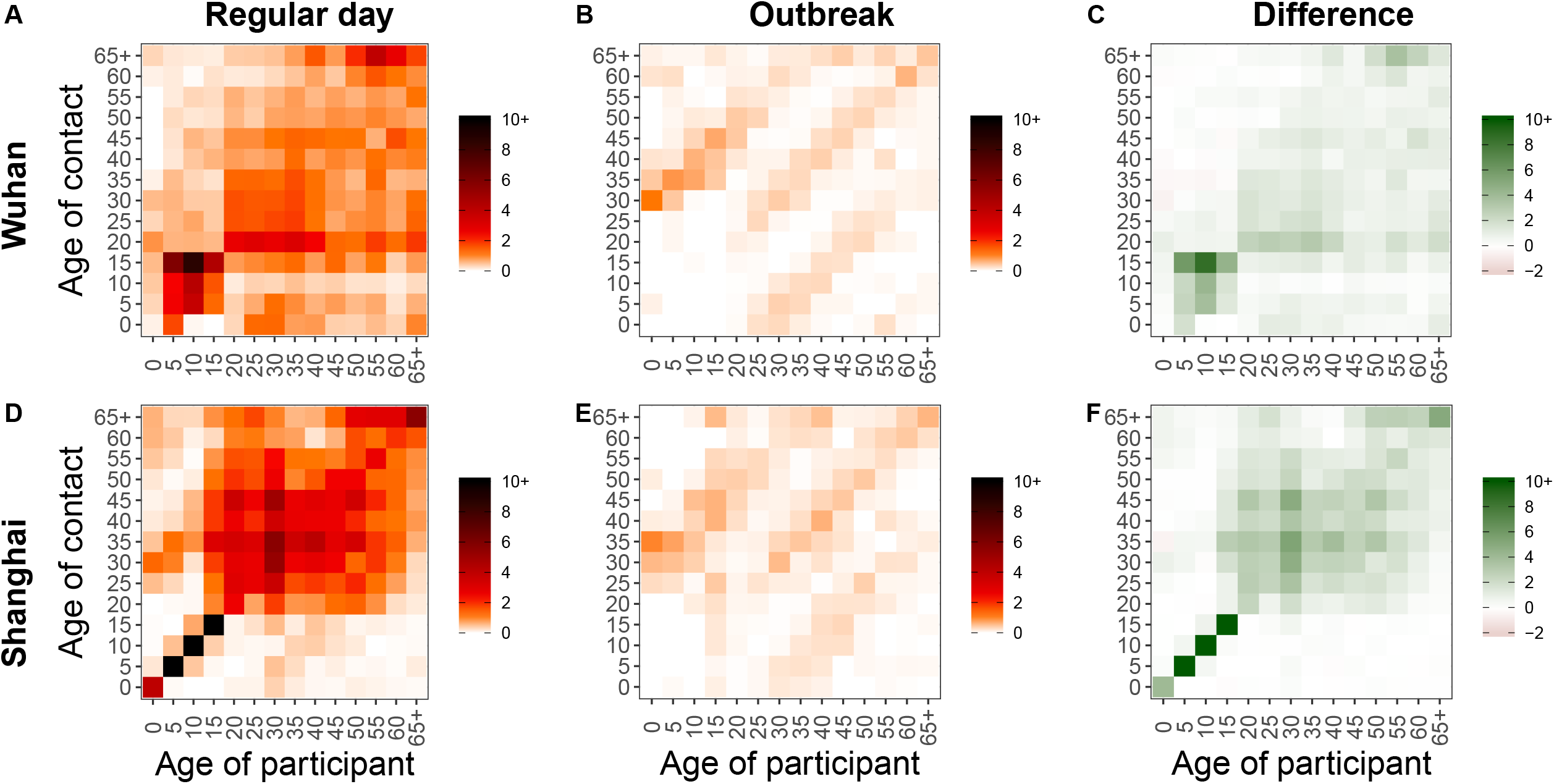
**A** Regular weekday contact matrix for Wuhan. An alternative visualization of this contact matrix is shown in Appendix (Sec.11). **B** Outbreak contact matrix for Wuhan. **C** Difference between the regular weekday contact matrix and the outbreak contact matrix in Wuhan. **D-F** Same as A-C, but for Shanghai.

To understand the interplay between social distancing interventions, changes in human mixing patterns, and outbreak dynamics, we also need to consider potential age differences in susceptibility to infection. This is currently a topic of debate, as little information on the age profile of asymptomatic cases is available(*7, 8*). To this aim, we analyzed contact tracing information from detailed epidemiological field investigations conducted by the Hunan CDC (Supplement). We calculated the age-specific relative risk of infection for close contacts of confirmed COVID-19 index cases. Briefly, all close contacts of COVID-19 cases reported in Hunan province were placed under medical observation for 14 days and were tested using real-time RT-PCR. Those who developed symptoms and tested positive were considered as symptomatic confirmed cases, while contacts who tested positive without exhibiting symptoms were considered as asymptomatic infections. The total of symptomatic and asymptomatic infections was used to estimate the relative susceptibility to infection by age. The ratio of symptomatic cases to total infections was used to estimate the relative probability of developing symptoms (see Supplementary Material). We found age differences in susceptibility to SARS-CoV2 infection, where young individuals (aged 0-14 years) had a lower risk of infection than individual ages 65 years and over (OR=0.41 (95%CI: 0.18-0.93), p-value=0.026). There was a weak non-significant trend towards lower susceptibility in middle-aged adults, relative to seniors (OR=0.76, 95%CI: 0.46-1.24, Tab. 2). These findings are in contrast with a previous study in Shenzhen, where susceptibility to infection did not change with age (*7*). Moreover, we found that the relative probability of developing symptoms also increased with age, however the difference was not statistically significant (Tab. 2).

Based on the estimated age-specific mixing patterns and susceptibility to COVID-19 infection, we developed a SIR model of SARS-CoV-2 transmission and tested the impact of social distancing measures on disease dynamics. In the model, the population is divided into three epidemiological categories: susceptible, infectious, and removed (either recovered or deceased individuals), stratified by 14 age groups. Susceptible individuals can become infectious after contact with an infectious individual according to the estimated age-specific susceptibility to infection. Because we did not see age differences in the probability of developing symptoms upon infection, we assumed equal infectivity across all age groups. The rate at which contacts occur is determined by the estimated mixing patterns in each age group. A key parameter regulating the dynamics of the model is the basic reproduction number (R_0_), which corresponds to the average number of secondary cases generated by a primary case in a fully susceptible population. The mean time interval between two consecutive generations of cases was taken to be 5.1 days (*2*). Details are reported in the Supplementary Materials.

For baseline R_0_ values in the range 2.0-3.5 associated with a regular weekday contact patterns (corresponding to the early phase of COVID-19 spread in Wuhan (*9-15*)), we find that the profound alteration of mixing patterns of the magnitude observed in Wuhan and Shanghai leads to a drastic decrease in R_0_. When we consider contact matrices representing the outbreak period, keeping the same baseline disease transmissibility as the pre-intervention period, the reproductive number drops well below the epidemic threshold both in Wuhan and Shanghai (Fig. 2A). In an uncontrolled epidemic (without intervention measure, travel restriction, or spontaneous behavioral response of the population), we estimate the mean infection attack rate to be in the range 64%-92% after a year of SARS-CoV-2 circulation, with slight variation between Wuhan and Shanghai (variations of about 5%-12%, Fig. 2B). On the other hand, if we consider a scenario where social distancing measures are implemented early on, as the new virus emerges, the estimated R_0_ remains under the threshold and thus the epidemic cannot take off in either location. Furthermore, we estimate that the magnitude of interventions implemented in Wuhan and Shanghai would have been enough to bring the reproduction number below 1.0 for baseline R_0_ up to ∼7 in Wuhan and ∼11.5 in Shanghai (Fig. 2A). We also conduct sensitivity analyses on assumptions about the susceptibility profile of infections, since there is still uncertainty about this parameter (see Supplementary Material). Our conclusions are robust to assuming equal susceptibility in all age groups.

**Figure 2.**
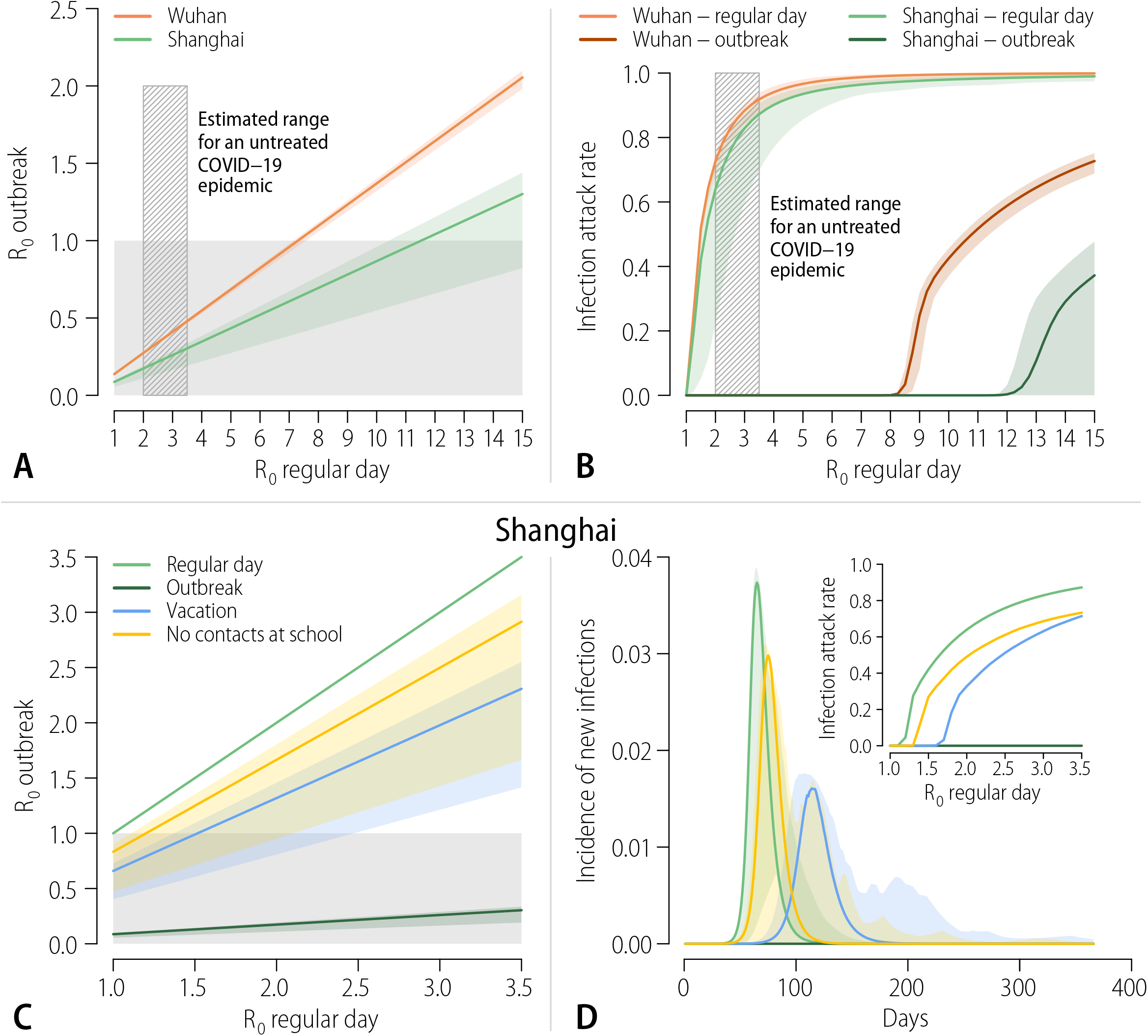
**A** Estimated R_0_ obtained with the outbreak contact matrix, as a function of baseline R_0_ derived from the regular weekday contact matrix, where the transmission rate is held constant. The solid light grey rectangle represents the region where the R_0_ in the outbreak situation is under the epidemic threshold. The shaded dashed black rectangle represents a plausible range of R_0_ values in the early phase of the COVID-19 epidemic, as estimated in the literature. **B** Infection attack rate a year after the initial case of COVID-19 as a function of baseline R_0,_ derived from the regular weekday contact matrix. The corresponding value of R_0_ for the outbreak situation can be seen in panel A. The shaded colored areas represent 95%CI. **C** As A, but for Shanghai only and including two scenarios of contact patterns reduction: (i) during vacations (Vacation) and (ii) during regular weekdays, after removing all contacts occurring in school setting (No schools). **D** Daily incidence of new infections for the four scenarios presented in panel C (median and 95%CI). The inset shows the mean infection attack rate after a year.

Finally, armed with a carefully calibrated model, we turn to assess the impact of a preemptive mass school closure. We used the same estimates of relative susceptibility to infection by age as in our main analysis (see Supplementary Material for a sensitivity analysis assuming equal susceptibility to infection). We considered two different contact pattern scenarios, based on data from Shanghai: contacts estimated during vacations period (*6*) and contacts estimated during regular weekdays, after all contacts occurring in school settings have been removed (*6*). Either one of these scenarios represent a simplification of a school closure strategy as during vacations children can still attend additional education, while removing all contacts in the school setting does not take into account that, for instance, parents may need to leave work to take care of children. We estimated that limiting contact patterns to those observed during vacations would interrupt transmission for baseline R_0_ up to 1.5 (Fig. 2C). Removing all school contacts would do the same for baseline R_0_ up to 1.2. If we apply these interventions to a COVID-19 scenario, assuming a baseline R0 of 2 - 3.5, we can achieve a noticeable decrease in infection attack rate and peak incidence, and a delay in the epidemic, but transmission is not interrupted (Fig. 2D). For instance, for baseline R_0_=2.5 and assuming a vacation mixing pattern, the peak daily incidence is reduced by about 57%. In the corresponding scenario where school contacts are removed, we estimate a reduction of about 20%. Overall, school-based policies are not sufficient to entirely prevent a COVID-19 outbreak but they can have a significant impact on disease dynamics, and hence on hospital surge capacity.

This study suffers from several limitations. First, the estimated mixing patterns for a regular weekday in Wuhan may be affected by recall bias since contacts were assessed retrospectively. For Shanghai, we relied on a survey conducted in December 2017 - May 2018, using the same design as the one conducted during the outbreak period, thus avoiding recall bias. It is also important to note that changes in contact patterns were measured in a context where social distancing was applied together with rapid isolation of infected individuals (including suspected cases) and quarantine of close contacts for 14 days. Only a small portion of the population in the two study sites was affected by contact tracing and quarantine, thus having little to no effect on the average contact patterns of the general population. However, in reconstructing the observed epidemics in Wuhan and Shanghai, it is not possible to separate the effects of case-based strategies from population wide social distancing. In our simulation model, we estimated the effect of social distancing alone; combining social distancing and case-based interventions would have a synergistic effect to further reduce transmission. Further, our estimates of age differences in susceptibility to infection and probability of developing symptoms are based on active testing of contacts of 57 primary confirmed cases. These data suffer from the usual difficulties inherent to identifying epidemiological links and index cases. Seroepidemiology studies are currently lacking but will be essential to fully resolve the population susceptibility profile of COVID-19.

While the age patterns of contacts were not significantly different between the two study locations during the COVID-19 outbreak period, these patterns may not be fully representative of other locations in China and abroad, where social distancing measures may differ. Modeling results may possibly be underestimating the effect of social distancing interventions as our results account for a decreased number of contacts but ignore the impact of increased awareness of the population, which may have also affected the type of social interactions (e.g., increased distance between individuals while in contact, or use of face mask(*16, 17*)). Finally, our school closure simulations are not meant to formulate a full intervention strategy, which would require identification of epidemic triggers to initiate closures and evaluation of different durations of intervention (*5*). Nonetheless, our modeling exercise provides an indication of the possible impact of a nation-wide preemptive strategy on the infection attack rate and peak incidence. To generalize these findings to other contexts, location-specific age-mixing patterns and population structures should be considered.

Our study provides evidence that the interventions put in place in Wuhan and Shanghai, and the resulting changes in human behavior, drastically decreased daily contacts, essentially reducing them to household interactions. Assuming the same scale of contact-distancing measures were to be put in place in other locations, human mixing patterns could be captured by data on within-household contacts, which are available for several countries around the world (*4-6, 18-20*). Further research should concentrate on refining age-specific estimates of susceptibility to infection, disease, and infectivity, which are instrumental to evaluating the impact of school- and work-based control strategies currently under consideration worldwide (*21, 22*).

## Data Availability

All data is available in the main text or the supplementary materials.

---

**Table 1.** Number of contacts by demographic characteristics and location, Wuhan and Shanghai.

| Characteristics | Wuhan |  |  |  |  | Shanghai |  |  |  |  |
| --- | --- | --- | --- | --- | --- | --- | --- | --- | --- | --- |
|  | Regular weekday |  | COVID-19 Outbreak |  | Difference | Regular weekday |  | COVID-19 Outbreak |  | Difference |
|  | N (%)<sup>a</sup> | Mean (95% CI) | N (%)<sup>a</sup> | Mean (95% CI) |  | N (%) | Mean (95% CI) | N (%) | Mean (95% CI) |  |
| Overall | 624 (100.0) | 14.6 (13, 16.3) | 627 (100.0) | 2.0 (1.9, 2.1) | 12.6<sup>***</sup> | 543 (100.0) | 20.6 (19, 22.3) | 557 (100.0) | 2.3 (1.9, 2.7) | 18.3<sup>***</sup> |
| Sex |  |  |  |  |  |  |  |  |  |  |
| Male | 300 (48.1) | 14.5 (12.2, 16.9) | 301 (48) | 1.8 (1.7, 2) | 12.6<sup>***</sup> | 264 (48.6) | 21.3 (18.8, 23.8) | 286 (51.3) | 2.1 (1.8, 2.3) | 19.2<sup>***</sup> |
| Female | 324 (51.9) | 14.7 (12.4, 17) | 326 (52) | 2.1 (2, 2.3) | 12.5<sup>***</sup> | 279 (51.4) | 20 (17.8, 22.2) | 271 (48.7) | 2.6 (1.8, 3.3) | 17.4<sup>***</sup> |
| Age group |  |  |  |  |  |  |  |  |  |  |
| 0-6 y | 12 (1.9) | 8.6 (0, 17.7) | 12 (1.9) | 2.2 (1.5, 2.8) | 6.4 | 51 (9.4) | 11.5 (8.1, 14.9) | 14 (2.5) | 1.9 (1.7, 2.2) | 9.6<sup>***</sup> |
| 7-19 y | 79 (12.7) | 16.2 (12.5, 19.8) | 79 (12.6) | 2.1 (2, 2.2) | 14.1<sup>***</sup> | 83 (15.3) | 33 (29.2, 36.9) | 55 (9.9) | 2.6 (1.7, 3.4) | 30.5<sup>***</sup> |
| 20-39 y | 254 (40.7) | 15.3 (12.6, 17.9) | 256 (40.8) | 2.1 (1.9, 2.2) | 13.2<sup>***</sup> | 132 (24.3) | 24.6 (20.8, 28.5) | 254 (45.6) | 2.2 (2, 2.5) | 22.4<sup>***</sup> |
| 40-59 y | 221 (35.4) | 13.8 (11, 16.6) | 220 (35.1) | 2 (1.8, 2.2) | 11.8<sup>***</sup> | 126 (23.2) | 20.4 (17.2, 23.7) | 160 (28.7) | 2.8 (1.6, 4.1) | 17.6<sup>***</sup> |
| >59 y | 58 (9.3) | 13.9 (7.3, 20.5) | 60 (9.6) | 1.4 (1.2, 1.7) | 11.6<sup>**</sup> | 151 (27.8) | 13.5 (10.9, 16.1) | 74 (13.3) | 1.6 (1.3, 1.8) | 11.9<sup>***</sup> |
| Type of profession |  |  |  |  |  |  |  |  |  |  |
| Pre-school | 12 (1.9) | 8.6 (0, 17.7) | 12 (1.9) | 2.2 (1.5, 2.8) | 6.4 | 43 (7.9) | 10 (6.4, 13.5) | 14 (2.5) | 1.9 (1.7, 2.2) | 8<sup>***</sup> |
| Student | 107 (17.1) | 14.6 (11.3, 18) | 107 (17.1) | 2.1 (2, 2.3) | 12.5<sup>***</sup> | 100 (18.5) | 31.1 (27.7, 34.5) | 71 (12.7) | 2.5 (1.8, 3.1) | 28.6<sup>***</sup> |
| Employed | 391 (62.7) | 15.4 (13.3, 17.5) | 390 (62.2) | 2.1 (1.9, 2.2) | 13.2<sup>***</sup> | 236 (43.6) | 23.9 (21.2, 26.6) | 354 (63.6) | 2.5 (2, 3.1) | 21.4<sup>***</sup> |
| Unemployed | 30 (4.8) | 14.1 (4.2, 24) | 31 (4.9) | 1.8 (1.3, 2.4) | 12.2<sup>*</sup> | 10 (1.8) | 12.4 (0.8, 24) | 24 (4.3) | 1.8 (1.3, 2.4) | 10.6 |
| Retired | 84 (13.5) | 12.1 (7, 17.2) | 87 (13.9) | 1.5 (1.3, 1.7) | 10.6<sup>***</sup> | 152 (28.1) | 12 (9.7, 14.3) | 94 (16.9) | 1.6 (1.3, 1.8) | 10.4<sup>***</sup> |
| Household size |  |  |  |  |  |  |  |  |  |  |
| 1-2 | 118 (18.9) | 11.8 (7.8, 15.8) | 121 (19.3) | 0.9 (0.6, 1.2) | 10.9<sup>***</sup> | 166 (30.6) | 15.6 (12.8, 18.4) | 199 (35.7) | 1.1 (0.8, 1.3) | 14.6<sup>***</sup> |
| 3-4 | 415 (66.5) | 13.9 (12.1, 15.7) | 415 (66.2) | 2 (2, 2.1) | 11.9<sup>***</sup> | 306 (56.4) | 21.8 (19.8, 23.9) | 294 (52.8) | 2.4 (2.3, 2.5) | 19.4<sup>***</sup> |
| >4 | 91 (14.6) | 21.5 (15.8, 27.1) | 91 (14.5) | 3.2 (2.9, 3.4) | 17.8<sup>***</sup> | 71 (13.1) | 27 (21.3, 32.7) | 64 (11.5) | 5.9 (2.8, 8.9) | 21.1<sup>***</sup> |

<sup>a</sup>Can differ from total sample size (n=636) as it also includes participants who had not recorded contacts during regular weekdays or during the COVID-19 outbreak. Note that reduced denominators indicate missing data. Percentages may not total 100 because of rounding.
\*p<0.05, \*\*p<0.01, \*\*\*p<0.001.

**Table 2.**
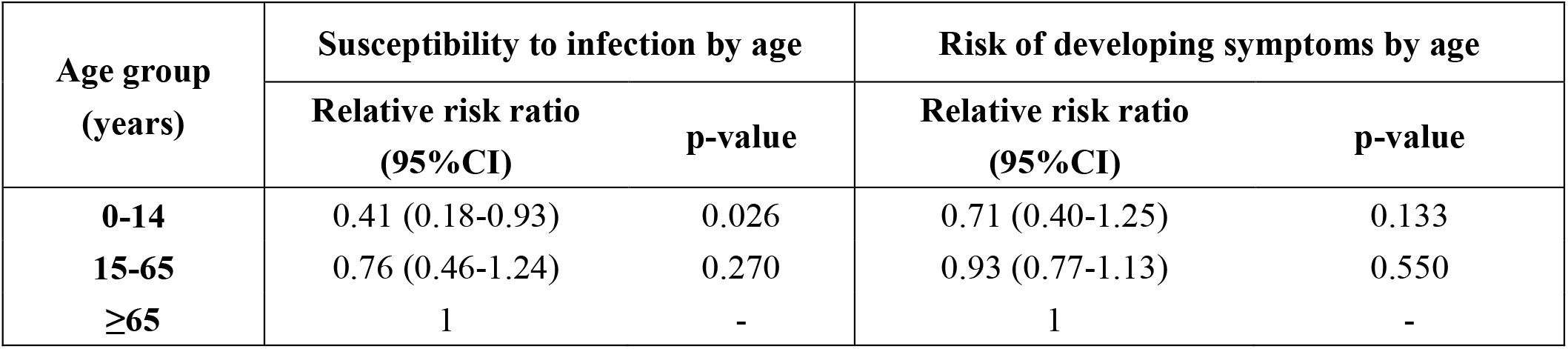
Age-specific estimates of relative susceptibility to infection and relative probability of developing symptoms with COVID-19.

## Notes

### Competing Interest Statement

A.V. has received funding from Metabiota Inc. H.Y. has received research funding from Sanofi Pasteur, GlaxoSmithKline, Yichang HEC Changjiang Pharmaceutical Company, and Shanghai Roche Pharmaceutical Company. This article does not necessarily represent the views of the NIH or the US government.

### Funding Statement

H.Y. acknowledges financial support from the National Science Fund for Distinguished Young Scholars (No. 81525023), Key Emergency Project of Shanghai Science and Technology Committee (No. 20411950100), National Science and Technology Major Project of China (No. 2018ZX10201001-010, No. 2018ZX10713001-007, No. 2017ZX10103009-005). The funder of the study had no role in study design, data collection, data analysis, data interpretation, or writing of the report. S.M. and M.A. acknowledge financial support from the European Commission H2020 MOOD project.

